# Primary Care Post-COVID syndrome Diagnosis and Referral Coding

**DOI:** 10.1101/2023.05.23.23289798

**Authors:** Robert Willans, Gail Allsopp, Pall Jonsson, Fiona Glen, John Macleod, Yinghui Wei, Felix Greaves, Sebastian Bacon, Amir Mehrkar, Alex Walker, Brian MacKenna, Louis Fisher, Ben Goldacre, The OpenSAFELY Collaborative, The CONVALESCENCE Collaborative

## Abstract

**Introduction:** Guidelines for diagnosing and managing Post-COVID syndrome have been rapidly developed. Consistency of the application of these guidelines in primary care is unknown. Electronic health records provide an opportunity to review the use of codes relating to Post-COVID syndrome. This paper explores the use of primary care records as a surrogate uptake measure for NICE’s rapid guideline “managing the long-term effects of COVID-19” by measuring the use of Post-COVID syndrome diagnosis and referral codes in the pathway.

**Method:** With the approval of NHS England we used routine clinical data from the OpenSafely-EMIS/-TPP platforms. Counts of Post-COVID syndrome diagnosis and referral codes were generated from a cohort of all adults, establishing numbers of diagnoses and referrals following diagnosis. The relationship between Post-COVID syndrome diagnosis and referral codes was explored with reference to NICE’s rapid guideline.

**Results:** Of over 45 million patients, 69,220 (0.15%) had a Post-COVID syndrome diagnostic code, and 67,741 (0.15%) had a referral code. 78% of referral codes did not have an associated diagnosis code. 79% of diagnosis codes had no subsequent referral code. Only 18,633 (0.04%) had both. There were higher rates of both diagnosis and referral in those who were more deprived, female and some ethnic groups.

**Discussion:** This study demonstrates variation in diagnosis and referral coding rates for Post-COVID syndrome across different patient groups. The results, with limited crossover of referral and diagnostic codes, suggest only one type of code is usually recorded. Recording one code limits the use of routine data for monitoring Post-COVID syndrome diagnosis and management, but suggests several areas for improvement in coding. Post-COVID syndrome coding, particularly diagnosis coding, needs to improve before administrators and researchers can use it to evaluate care pathways.

## Introduction

The long-term harmful consequences of Covid are uncertain, but given the scale of population level Covid infection, the potential harms are of substantial interest to individuals, health services and national economies.

Population level surveys have estimated that 3.5% of the population in England experienced self-reported symptoms of Post-COVID syndrome in September 2022 (symptoms persisting more than 4 weeks after the first COVID-19 infection) [1].

NICE has moved to rapidly develop guidance for the diagnosis and management of Post-COVID syndrome using emerging case definitions. NICE’s guideline “managing the long-term effects of COVID-19”[2] sets out several recommendations for identifying, assessing and managing the long-term effects of COVID-19, many of which can be expressed in terms of clinical codes.

Electronic health records (EHRs) have the potential to measure uptake of best practice recommendations produced by guideline-writing organisations such as NICE. Measuring uptake is a key step in disseminating and promoting new guidance. Where the uptake of guidance lags, is inconsistent, or there is a differential uptake in different populations and groups, there may be opportunities to improve practitioner knowledge and consistency, optimise service provision, and drive continuous improvement in adoption.

Measuring NICE guidance uptake is particularly important for new conditions such as Post-COVID syndrome. New conditions have no previous information on uptake, services are usually being developed, and practitioners may have a less clear understanding of emerging standards of care.

Measuring compliance through EHRs requires recommendations to be aligned to codeable events in the health record and for coding to be as comprehensive as possible. For primary care of Post-COVID syndrome, the most measurable recommendation concerns when suspected long-covid cases are referred to secondary care. It is important to establish how clinical coding in this area is used in practice and whether the coding quality can support inferences about uptake and consistency with the associated guideline recommendations. Of particular importance for measuring uptake is whether diagnosis codes are being recorded as expected. Without a level of confidence in diagnosis codes, it is hard to define a cohort of Post-COVID syndrome patients to effectively measure guidance implementation.

Previous work in this area has been carried out by Walker et al.[3] who found primary care coding rates of Post-COVID syndrome were lower than other estimates of self-reported prevalence. Usage of codes also varied by the software system used.

The use of these diagnosis codes is a prerequisite for collecting real world information on prevalence, symptoms, and treatments for Post-COVID syndrome. Without this information, those planning services may struggle to design services appropriately, especially where multidisciplinary provision is needed, and service requirements are uncertain [4].

More broadly, NICE is interested in how their guidance can be made as ‘computable’ as possible, allowing automated, continuous, and close to real time measurement of uptake and variation. Authors have started to theorise the process of creating ‘rapid learning systems’ which relate available data to NICE’s guidance to develop machine-interpretable knowledge [5,6].

We consequently set out to quantify how frequently diagnosis and referral codes for the long-term effects of COVID-19 were being used - and how this varied - as a component measure of compliance with NICE’s long-term effects of COVID syndrome guideline.

## Method

### Study design & data sources

A retrospective cohort analysis was carried out to examine the use of Post-COVID syndrome diagnosis and referral codes in primary care. Recordings of diagnostic and referral codes were obtained from the OpenSAFELY TPP and EMIS databases. These databases comprise all patients in England whose GP practice use either TPP (SystmOne) or EMIS as an EHR. All data were linked, stored and analysed securely within the OpenSAFELY platform: https://opensafely.org/. Data include pseudonymised data such as coded diagnoses, medications and physiological parameters. No free text data are included. All code is shared openly for review and re-use under MIT open license https://github.com/opensafely/Post_covid_nice_compliance. Detailed pseudonymised patient data is potentially re-identifiable and therefore not shared.

After extracting population data, a descriptive analysis of diagnoses and referrals was carried out as follows: Firstly, counts of diagnostic and referral codes were extracted from the database using queries and the codelist in the Appendix. Secondly, the relationship between diagnosis and referral codes was investigated to identify if referral rates could be computed and if any inferences about uptake of NICE guidance could be made. We investigated the correlation between diagnosis and referral codes to investigate the feasibility of calculating representative referral rates and adherence to NICE’s “managing the long-term effects of COVID-19” guideline. Finally, subdivisions of each diagnosis and referral code were investigated to examine how the number of diagnosis or referral codes differed according to demographic characteristics.

### Study population

All adults (aged 18 or over) registered with a general practice as of 1st February 2022 were included, with data taken for the preceding 36 months. Those with incomplete age or gender fields were excluded.

### Outcomes and clinical coding

We use the presence of diagnosis and referral codes in the primary care record as outcomes, stratified by demographic variables constructed from age, sex, geographical region, ethnicity, and socioeconomic deprivation. Socioeconomic deprivation was based on Index of Multiple Deprivation (IMD) quintiles.

Diagnosis and referral codes used are taken from the new SNOMED Codes introduced in December 2020 and implemented in 2021. The new codes record diagnoses, referrals, and treatments of Post-COVID syndrome. These SNOMED codes included two diagnosis codes, differentiating between diagnoses of “Ongoing symptomatic COVID-19” and “Post-COVID-19 syndrome”. Ongoing symptomatic COVID-19 is defined as “signs and symptoms of COVID-19 from 4 weeks up to 12 weeks”, with “Post-COVID-19 syndrome” defined as “signs and symptoms that…continue for more than 12 weeks”. Three main intervention codes were defined, one for a direction to the Your COVID Recovery (“YCR”) public website, one for a formal referral to the associated online rehabilitation platform for patients with Post-COVID-19 syndrome, and one code for a referral to Post-COVID-19 clinics.

### Statistical methods

Counts and relative frequencies for each code were obtained, with confidence intervals for rates computed as a normal approximation to the Poisson distribution.

### Software and reproducibility

Data management was performed using Python 3, with analysis carried out using R. Code for data management and analysis, as well as codelists, are archived online at https://github.com/opensafely/Post_covid_nice_compliance.

## Results

Characteristics of the cohort are set out in Table 1. The total number of people included in the cohort was 45,782,618, 19,017,433 from TPP and 26,765,185 from EMIS. This cohort represents roughly 93% of the English adult patient population. 24% (10,761,068/45,782,618) of ethnicity codes were unknown or not present, as were 1% (623,198/45,782,618) of IMD quintiles generated by postcode.

**Table 1:**
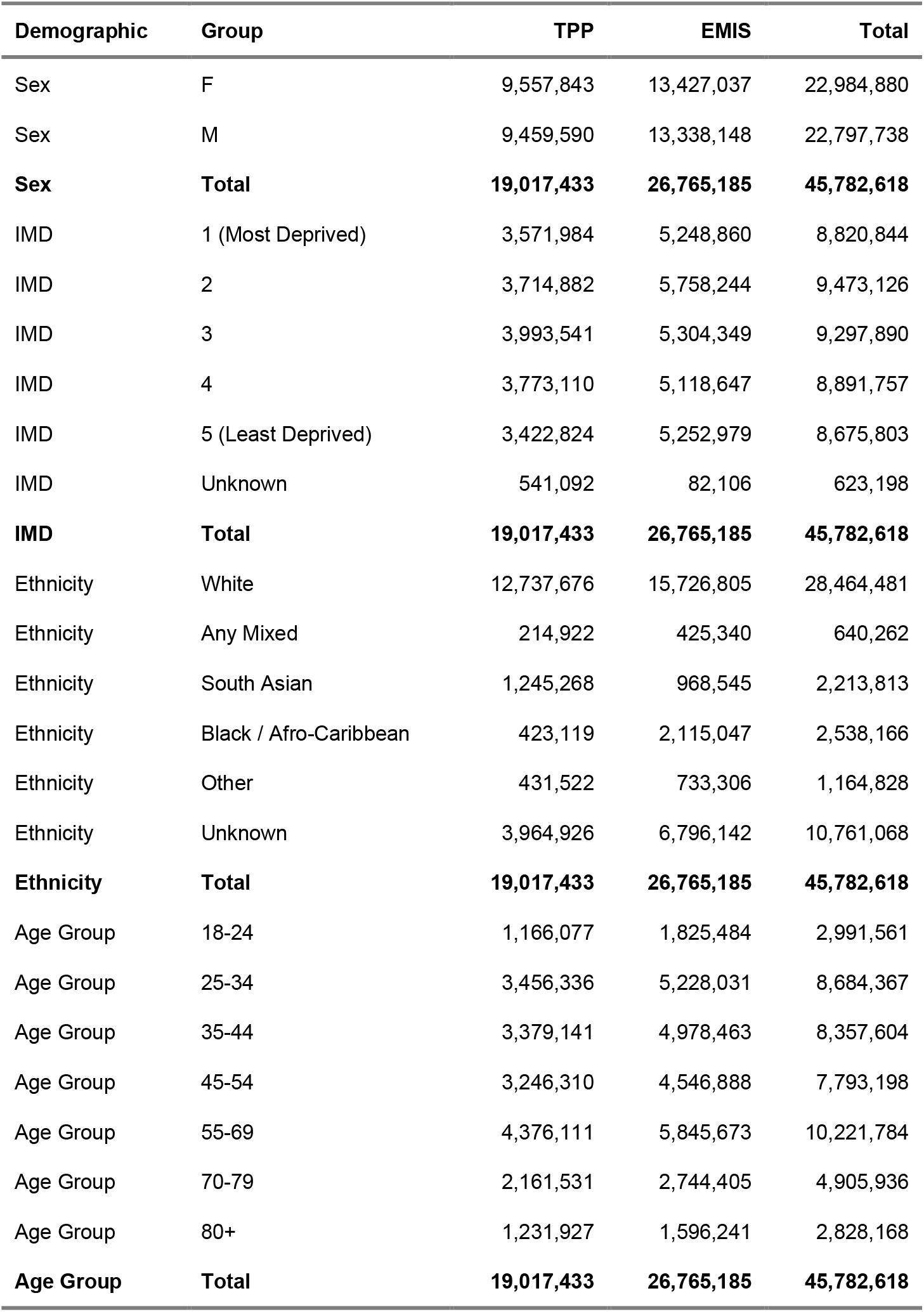
Patient Counts by Demographic Group

### Association between diagnosis and referral codes

78% (67,741/86,374) of those with referral codes to post-covid clinics had no recorded ongoing symptomatic COVID-19 or Post-COVID-19 diagnosis code (table 2). Similarly, 79% (69,220/87,853) of diagnosis codes do not have an associated referral or signpost code to further treatment. Given the observed low frequency and lack of correlation between diagnosis and referral codes, detailed referral rates were not calculated.

**Table 2:** Association between Diagnosis Codes and Referral Codes

| **Diag** | **No Referral Code** | **Referral Code** | **Total** |
| --- | --- | --- | --- |
| No Diagnosis Code | 45,627,024 | 67,741 | **45,694,765** |
| Diagnosis Code | 69,220 | 18,633 | **87,853** |
| **Total** | **45,696,244** | **86,374** | **45,782,618** |

### Diagnosis and referral codes through time

The frequency of Post-COVID syndrome diagnosis and referral codes has not substantially increased with time (Figure 2). Usage of the codes increased after implementation but reached a plateau at a very low level of usage. There was a peak in the use of the “Signposting to Your COVID Recovery” code around January 2022.

**Figure 1:**
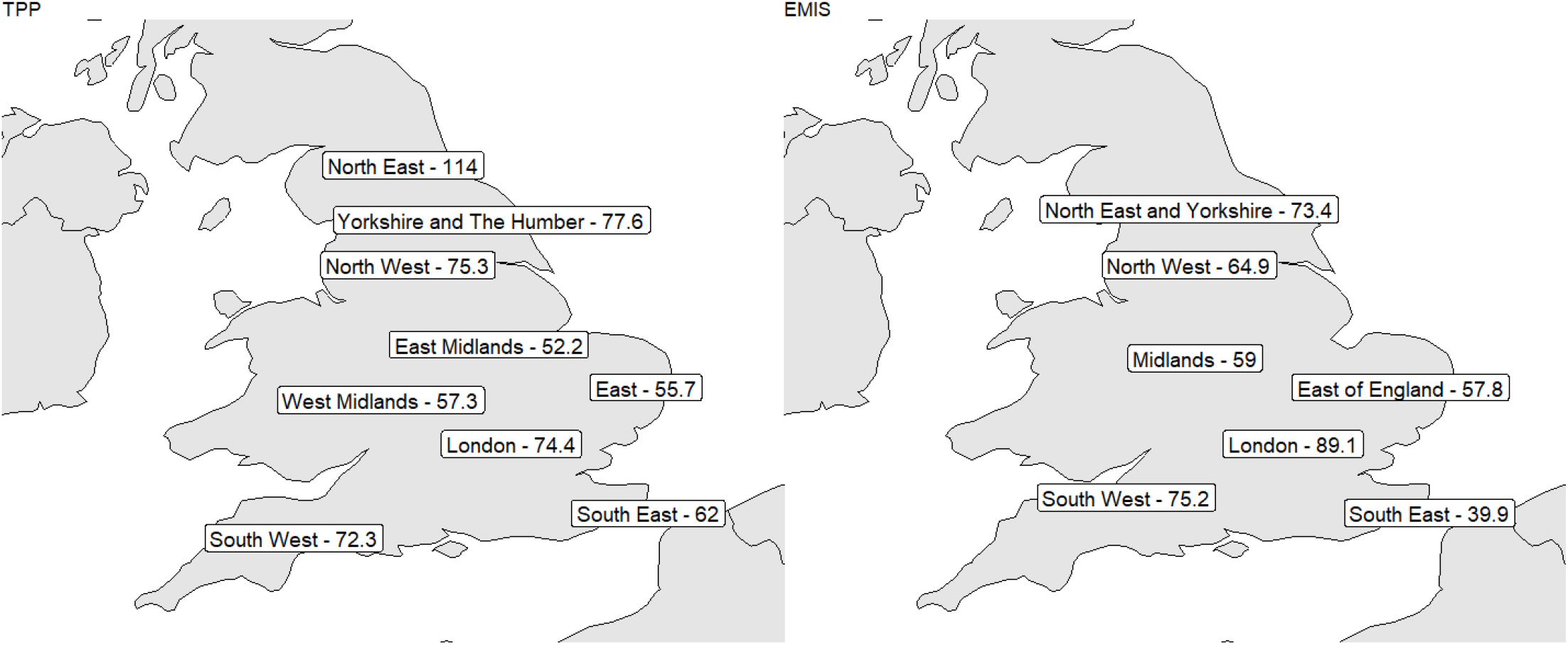
TPP and EMIS Post Covid Clinic referral rate per 100,000

**Figure 2:**
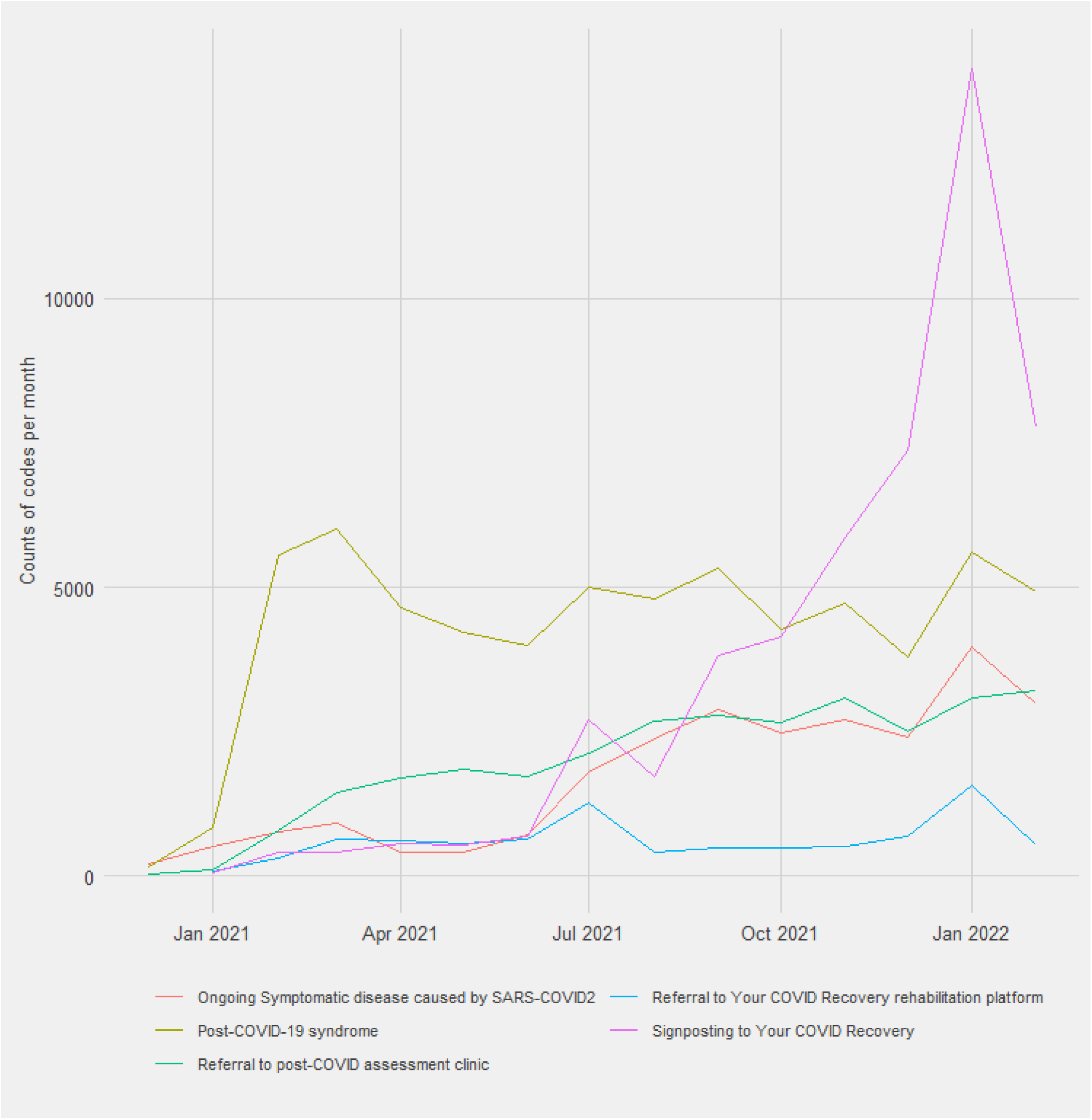
Code usage through time

### Demographic subgroups

Post-COVID syndrome diagnostic codes are more frequently recorded for female patients than males (Table 3), particularly the post-covid syndrome code. Female patients were 1.94 95% CI [1.89, 1.99] times as likely to have an ongoing symptomatic COVID-19 diagnosis code than male patients and 1.85 [1.82, 1.88] times more likely to have a Post-COVID-19 syndrome diagnosis code. Referrals also showed a similar pattern with female patients 1.66 [1.59, 1.73] times more likely to have a referral code to the YCR website program and 1.93 [1.88, 1.97] times as likely to receive a code indicating a referral to a Post-COVID-19 syndrome outpatient clinic.

**Table 3:** Demographic Split of ‘Long Covid’ Diagnosis and Referral Codes

| Demographic | Group | N | Ongoing symptomatic COVID-19 rate per 100000 | Post COVID-19 syndrome | Post-COVID-19 syndrome rate per 100000 | Refer YCR Website | Refer YCR Website rate per 100000 | Refer YCR Program | Refer YCR Program rate per 100000 | Refer Post-COVID-19 Clinic | Refer Post-COVID-19 Clinic rate per 100000 |
| --- | --- | --- | --- | --- | --- | --- | --- | --- | --- | --- | --- |
| Sex | F | 22984880 | 74.7 [73.6, 75.8] | 42366 | 184.3 [182.6, 186.1] | 29883 | 130 [128.5, 131.5] | 5530 | 24.1 [23.4, 24.7] | 19963 | 86.9 [85.6, 88.1] |
| Sex | M | 22797738 | 38.6 [37.8, 39.4] | 22779 | 99.9 [98.6, 101.2] | 22084 | 96.9 [95.6, 98.1] | 3303 | 14.5 [14, 15] | 10285 | 45.1 [44.2, 46] |
| Sex | Total | 45782618 | 56.7 [56, 57.4] | 65145 | 142.3 [141.2, 143.4] | 51967 | 113.5 [112.5, 114.5] | 8833 | 19.3 [18.9, 19.7] | 30248 | 66.1 [65.3, 66.8] |
| IMD | 1 (Most Deprived) | 8820844 | 65.2 [63.5, 66.9] | 14497 | 164.3 [161.7, 167] | 9175 | 104 [101.9, 106.1] | 1555 | 17.6 [16.8, 18.5] | 6207 | 70.4 [68.6, 72.1] |
| IMD | 2 | 9473126 | 57.4 [55.9, 58.9] | 14587 | 154 [151.5, 156.5] | 9656 | 101.9 [99.9, 104] | 1647 | 17.4 [16.5, 18.2] | 6774 | 71.5 [69.8, 73.2] |
| IMD | 3 | 9297890 | 53.5 [52, 55] | 12535 | 134.8 [132.5, 137.2] | 10563 | 113.6 [111.4, 115.8] | * | * [*, *] | 6262 | 67.3 [65.7, 69] |
| IMD | 4 | 8891757 | 54.8 [53.2, 56.3] | 11945 | 134.3 [131.9, 136.7] | 11253 | 126.6 [124.2, 128.9] | 1862 | 20.9 [20, 21.9] | 5567 | 62.6 [61, 64.3] |
| IMD | 5 (Least Deprived) | 8675803 | 53 [51.5, 54.6] | 11199 | 129.1 [126.7, 131.5] | 10929 | 126 [123.6, 128.3] | 1666 | 19.2 [18.3, 20.1] | 4984 | 57.4 [55.9, 59] |
| IMD | Unknown | 623198 | 51.5 [45.9, 57.1] | 382 | 61.3 [55.1, 67.4] | 391 | 62.7 [56.5, 69] | * | * [*, *] | 454 | 72.9 [66.1, 79.6] |
| IMD | Total | 45782618 | 56.7 [56, 57.4] | 65145 | 142.3 [141.2, 143.4] | 51967 | 113.5 [112.5, 114.5] | 8833 | 19.3 [18.9, 19.7] | 30248 | 66.1 [65.3, 66.8] |
| Ethnicity | Any Mixed | 640262 | 63.4 [57.2, 69.6] | 1091 | 170.4 [160.3, 180.5] | 835 | 130.4 [121.6, 139.3] | 90 | 14.1 [11.2, 17] | 429 | 67 [60.7, 73.3] |
| Ethnicity | Black / Afro-Caribbean | 2538166 | 71.7 [68.4, 75] | 5899 | 232.4 [226.5, 238.3] | 2523 | 99.4 [95.5, 103.3] | 190 | 7.5 [6.4, 8.6] | 2008 | 79.1 [75.7, 82.6] |
| Ethnicity | South Asian | 2213813 | 54.7 [51.6, 57.8] | 2843 | 128.4 [123.7, 133.1] | 1650 | 74.5 [70.9, 78.1] | 343 | 15.5 [13.9, 17.1] | 1535 | 69.3 [65.9, 72.8] |
| Ethnicity | White | 28464481 | 59.3 [58.4, 60.2] | 41362 | 145.3 [143.9, 146.7] | 33613 | 118.1 [116.8, 119.3] | 6317 | 22.2 [21.6, 22.7] | 20453 | 71.9 [70.9, 72.8] |
| Ethnicity | Other | 1164828 | 34 [30.6, 37.3] | 1015 | 87.1 [81.8, 92.5] | 851 | 73.1 [68.1, 78] | 79 | 6.8 [5.3, 8.3] | 471 | 40.4 [36.8, 44.1] |
| Ethnicity | Unknown | 10761068 | 48.7 [47.4, 50.1] | 12935 | 120.2 [118.1, 122.3] | 12495 | 116.1 [114.1, 118.1] | 1814 | 16.9 [16.1, 17.6] | 5352 | 49.7 [48.4, 51.1] |
| Ethnicity | Total | 45782618 | 56.7 [56, 57.4] | 65145 | 142.3 [141.2, 143.4] | 51967 | 113.5 [112.5, 114.5] | 8833 | 19.3 [18.9, 19.7] | 30248 | 66.1 [65.3, 66.8] |
| Age Group | 18-24 | 2991561 | 31.2 [29.2, 33.2] | 1842 | 61.6 [58.8, 64.4] | 5055 | 169 [164.3, 173.6] | 350 | 11.7 [10.5, 12.9] | 905 | 30.3 [28.3, 32.2] |
| Age Group | 25-34 | 8684367 | 46.4 [45, 47.9] | 8958 | 103.2 [101, 105.3] | 13495 | 155.4 [152.8, 158] | 1371 | 15.8 [15, 16.6] | 3915 | 45.1 [43.7, 46.5] |
| Age Group | 35-44 | 8357604 | 77.3 [75.4, 79.2] | 14866 | 177.9 [175, 180.7] | 12367 | 148 [145.4, 150.6] | 2087 | 25 [23.9, 26] | 6756 | 80.8 [78.9, 82.8] |
| Age Group | 45-54 | 7793198 | 87.3 [85.2, 89.4] | 17616 | 226 [222.7, 229.4] | 10322 | 132.4 [129.9, 135] | 2227 | 28.6 [27.4, 29.8] | 8419 | 108 [105.7, 110.3] |
| Age Group | 55-69 | 10221784 | 58.7 [57.2, 60.2] | 18033 | 176.4 [173.8, 179] | 8469 | 82.9 [81.1, 84.6] | 2296 | 22.5 [21.5, 23.4] | 8449 | 82.7 [80.9, 84.4] |
| Age Group | 70-79 | 4905936 | 24.9 [23.5, 26.3] | 2950 | 60.1 [58, 62.3] | 1715 | 35 [33.3, 36.6] | 394 | 8 [7.2, 8.8] | 1397 | 28.5 [27, 30] |
| Age Group | 80+ | 2828168 | 18.1 [16.5, 19.7] | 880 | 31.1 [29.1, 33.2] | 544 | 19.2 [17.6, 20.9] | 108 | 3.8 [3.1, 4.5] | 407 | 14.4 [13, 15.8] |
| Age Group | Total | 45782618 | 56.7 [56, 57.4] | 65145 | 142.3 [141.2, 143.4] | 51967 | 113.5 [112.5, 114.5] | 8833 | 19.3 [18.9, 19.7] | 30248 | 66.1 [65.3, 66.8] |

| Demographic | Group | N | Ongoing<br>symptomatic<br>COVID-19 rate<br>per 100000 | Post<br>COVID-19<br>syndrome | Post-COVID-19 syndrome<br>rate per<br>100000 | Refer<br>YCR<br>Website | Refer YCR<br>Website<br>rate per<br>100000 | Refer<br>YCR<br>Program | Refer YCR<br>Program<br>rate per<br>100000 | Refer<br>Post-COVID-19<br>Clinic | Refer Post-COVID-19<br>Clinic rate<br>per 100000 |
| --- | --- | --- | --- | --- | --- | --- | --- | --- | --- | --- | --- |

\* - Values suppressed due to small numbers

Across IMD quintiles, Ongoing symptomatic COVID-19 diagnosis codes were recorded more often at higher levels of deprivation, 65.2 per 100,000 [63.5, 66.9] in the most deprived quintile and 53 per 100,000 [51.5, 54.6] in the least. Post-COVID-19 syndrome diagnosis codes also followed this pattern. The most deprived quintile recorded 164.3 Post-COVID-19 diagnosis codes per 100,000 [161.7, 167] amongst the most deprived quintile vs 129.1 per 100,000 [126.7, 131.5] in the least deprived. Less deprived quintiles were more likely to have a code recording either a signpost to the YCR website or the YCR program delivered through the website. The least deprived quintile had 126 codes for website signposts per 100000 [123.6, 128.3], compared to 104 per 100000 [101.9, 106.1] in the most deprived quintile. There were 19.2 per 100000 [18.3, 20.1] referral codes for the program in the least deprived quintile compared to 17.6 [16.8, 18.5] in the most deprived. People in more deprived areas were more likely to have a referral code for a Post-COVID-19 clinic, with referral codes amongst the most deprived quintile (70.4 per 100000 [68.6, 72.1]) higher than the least deprived (57.4 per 100000 [55.9, 59]).

Ongoing symptomatic COVID-19 diagnosis codes were observed slightly more often in Black ethnicities (71.7 per 100,000 [68.4, 75]) and mixed ethnicities (63.4 per 100,000 [57.2, 69.6]) than other groups, though overall ethnicity recording in the dataset is low, with 24% missing. Post-COVID-19 diagnosis codes followed a similar pattern. The rate for Black ethnicities (232.4 per 100,000 [226.5, 238.3]) and Mixed ethnicities (170.4 per 100,000 [160.3, 180.5] was higher than the overall rate of 142.3 per 100,000 [141.1, 143.4].

Ongoing symptomatic COVID-19 and Post-COVID-19 diagnosis code rates were higher with age, peaking in the 45-54 age group. Ongoing symptomatic disease code recording was 87.3 per 100000 [85.2, 89.4] in this age group, with Post-COVID-19 diagnosis code recordings being 226 per 100000 [222.7, 229.4]. 83% of Post-COVID-19 syndrome diagnosis codes were recorded among patients aged 35 and over. Younger age groups were more likely to receive a code for a signpost to the YCR website, with those aged 18-24 years having the highest rate of 169 per 100000 [164.3, 173.6]. Referral codes to the YCR programme and to post covid clinics were highest amongst the 45-54 age group, where diagnosis codes were highest.

Regional splits were difficult to compare as regional categories were not fully comparable between the two systems but appeared to show lower usage of referral codes in the East of England and higher usage in London, the North-East and the South-West.

### Recording of codes through pathways

There was limited overlap between those who received both a diagnosis and referral code (table 1). While a similar number of people had a diagnosis code (69,220) and a referral code (67,741), only 18,663 had both. This is only 16% of those who received either a diagnosis or referral code.

There was also variation in coding between the different EHR systems. Few patients with a diagnosis code for Ongoing symptomatic COVID-19 have subsequently recorded referral codes. In TPP (Figure 4), fewer than a third have a recorded signpost code for the YCR website, and fewer than a sixth receive a subsequent Post COVID-19 diagnosis code. In EMIS (Figure 5), a lower proportion of those with an Ongoing symptomatic COVID-19 code have a code indicating a signpost to the YCR website. However, the conversion rate from an Ongoing symptomatic COVID-19 code to a Post-COVID-19 Syndrome diagnosis code is similar to TPP.

**Figure 3:**
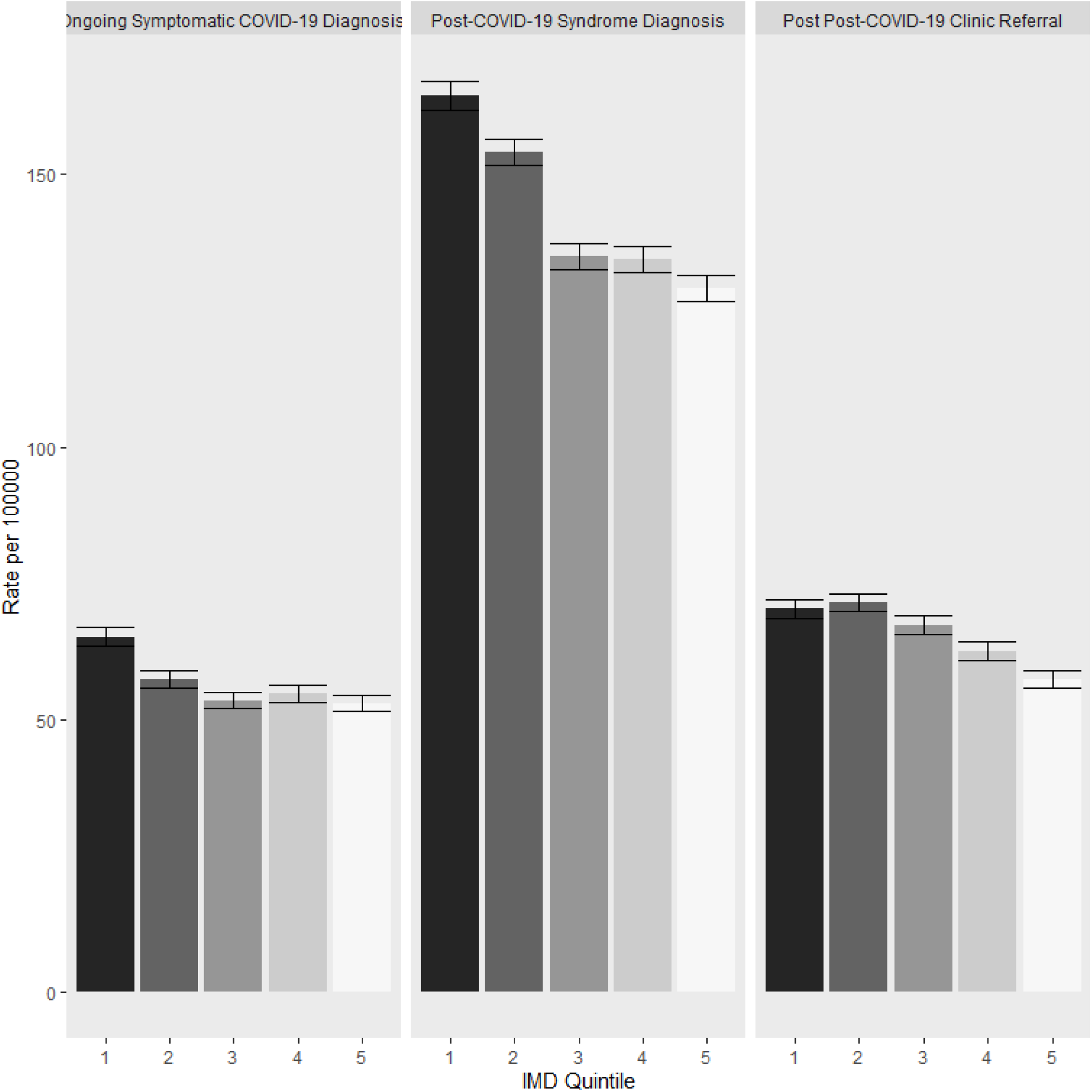
Diagnoses and Secondary Care Referrals by IMD Quintile (1 - Most Deprived, 5 - Least Deprived)

**Figure 4:**
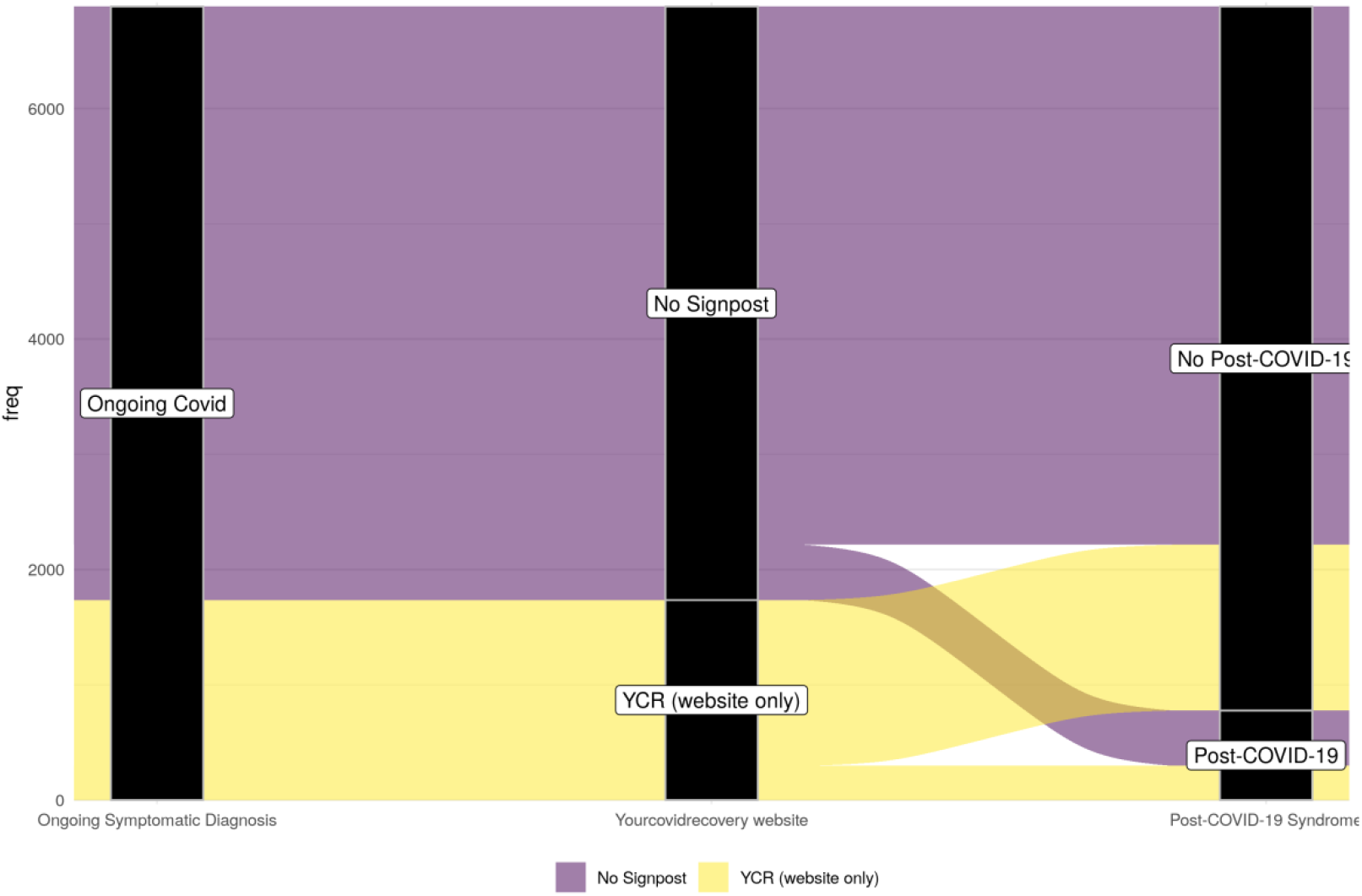
Destinations of those diagnosed with ongoing symptomatic covid - TPP

**Figure 5:**
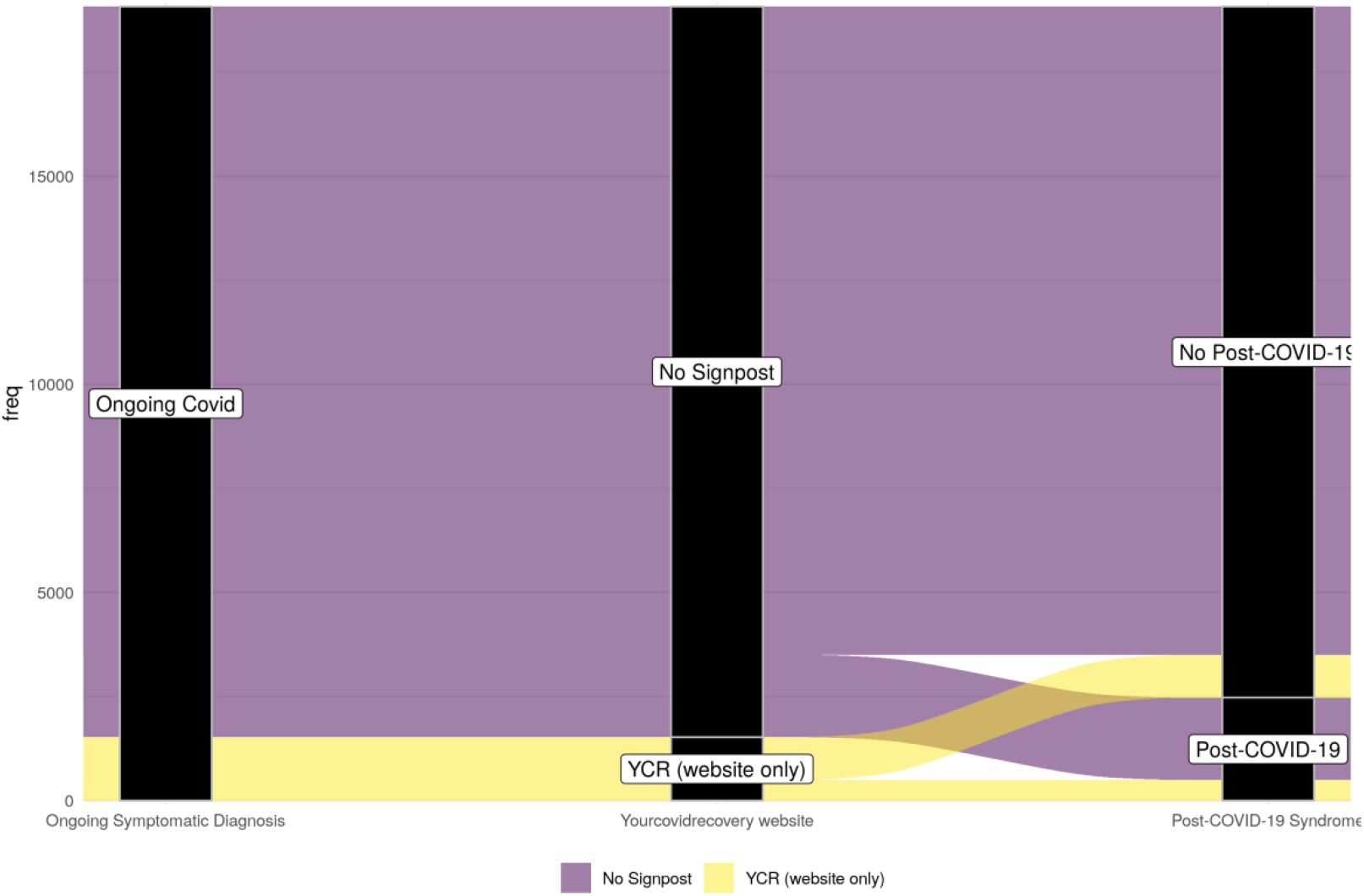
Destinations of those diagnosed with ongoing symptomatic covid - EMIS

Amongst those who do receive a Post-COVID-19 syndrome diagnosis code, few people have a referral code for the YCR programme. Conversion rates were lower in EMIS (Figure 7) than in TPP (Figure 6). Those who had a subsequent referral to a Post-COVID-19 service were more likely to have had a programme referral code.

**Figure 6:**
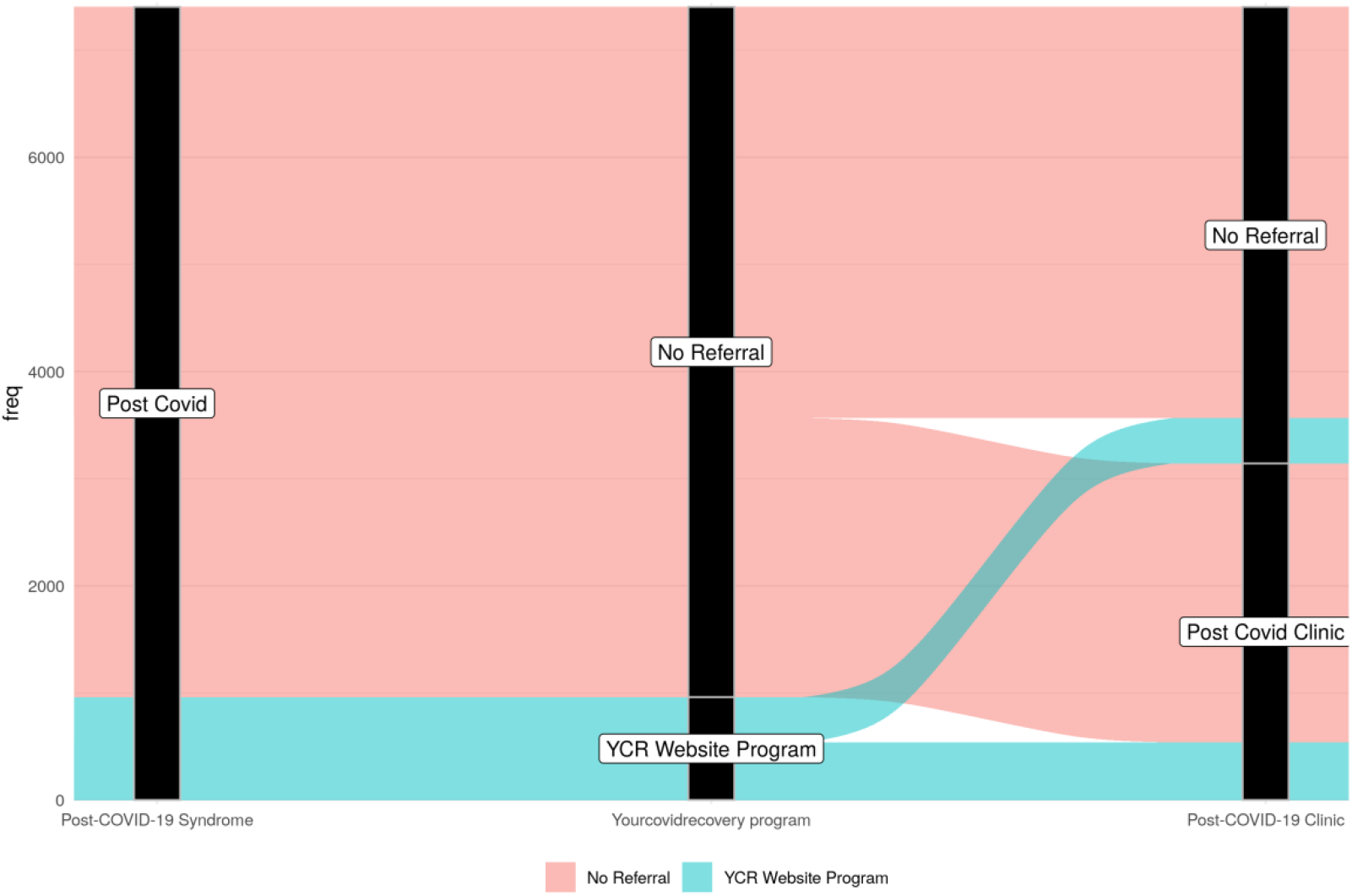
Destinations of those with a recorded Post-Covid Diagnosis - TPP

**Figure 7:**
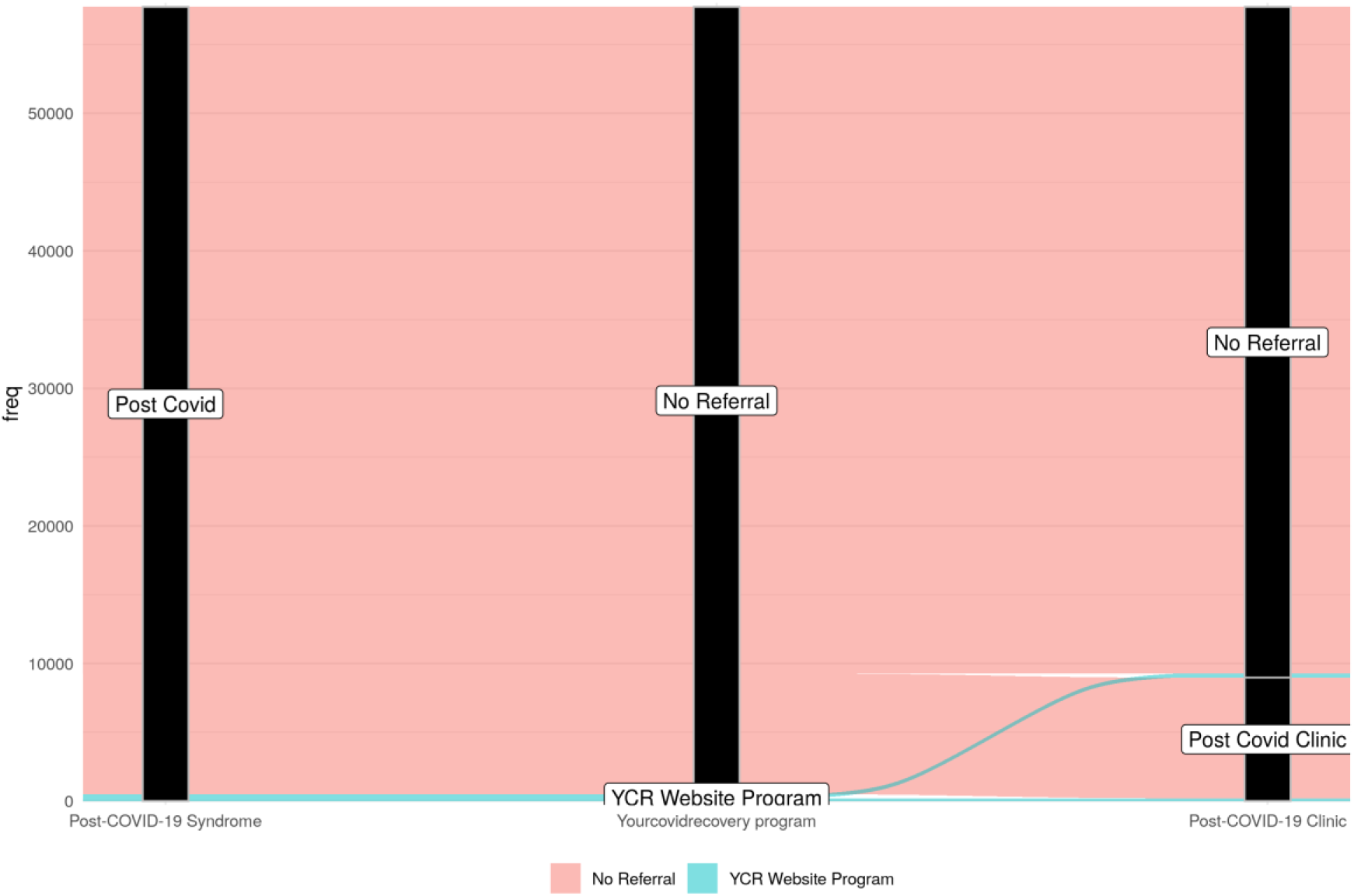
Destinations of those with a recorded Post-Covid Diagnosis – EMIS

## Discussion

### Summary

The results demonstrate that observable and potentially improvable patterns can be seen in the diagnosis and ongoing management of Post-COVID syndrome. Caution will need to be taken in interpretation due to the lack of completeness of recording of both diagnosis and referral in the EHR, as demonstrated by the lack of overlap between the populations with these codes recorded.

Overall use of clinical codes for Post-COVID syndrome in primary care is low. The total number of people with a diagnostic code (0.15% of the studied population) is markedly lower than those self-reporting ongoing symptoms in the ONS survey. While measuring different thresholds, these results suggest that either a large number of people with self-reported symptoms are not reporting these to the GP, or the GP is not coding them as Post-COVID syndrome.

The total number of people monthly with a referral code to Post COVID services is also lower than those reported in NHS England’s monthly data - 5,765 between 17th January and 13th February 2022 [7], suggesting not all referrals are captured in the EHRs. Variation in referral by various categories, including age, gender, and ethnicity, may represent variation in who the disease affects but may also reflect variation in service provision and access. Where diagnosis codes are recorded, rates amongst demographic subgroups appeared consistent with other findings. The observed gender disparity, with higher rates of female diagnosis, has already been noted in other analyses [8,9]. This observation gives us some confidence that the under-recording does not seriously skew the demographic information observed from this sample of codes. However, it is not possible to say whether variation in diagnosis code use is due to variation in true Post COVID-19 disease prevalence or variation in capturing and recording rates amongst the various sources of information.

Overall, this study confirms that coding depth of Post-COVID syndrome remains limited and has not markedly changed since the study by Walker et al.[3]. The differences in results between EHR systems, in terms of overall patterns of code use, variation in use, and overlap between diagnostic and referral codes suggests that some aspects of EHR system design (e.g. workflow, ergonomics or training) might be influencing coding patterns. This pattern merits further investigation.

### Measuring guideline compliance

To the best of our knowledge, there are no prior attempts to use clinical coding to assess compliance with NICE guidance on Post-COVID syndrome. The limitations of using routine EHR data are demonstrated here. The lack of consistency between diagnosis and referral codes limits the ability to measure referral patterns. Conditions such as Post-COVID syndrome, which have less clearly defined diagnostic criteria, may be less amenable to automated measurement than those with tightly defined diagnostic criteria (e.g. hypertension and diabetes)

NICE aspires to increase the computability of its guidance. Having guidance in a computable format, with contextual clinical codes, allows for the monitoring of uptake of that guidance. However, using EHRs to measure uptake is contingent on nominated codes being used in clinical settings. Attempts to move towards systemic use of EHRs for uptake monitoring are likely to be impeded if data quality, particularly diagnosis codes, are not used regularly. Other interested parties and researchers would also benefit from being able to use Post-COVID syndrome diagnostic codes to define cohorts for additional research.

More generally, there is a trend for the increasing use of administrative data in audits, including audits of NICE guidance, both in smaller studies [10] and larger national audits [11]. This study shows that consistency and depth of coding are important prerequisites for such approaches.

Further work on coding quality is needed to maximise the benefits of using EHR data for national healthcare management and planning. If diagnosis codes for Post-COVID syndrome remain under-recorded, it is harder to evaluate primary care pathways and care quality. A higher depth and consistency in primary care coding would allow researchers and national bodies to evaluate how well NICE guidance is implemented in practice. Without coding improvements, administrative data will likely be of limited audit use.

### Strengths and limitations

This study’s chief strength is the extensive coverage of England’s primary care records. Using two large-scale primary care records providers gives us close to universal coverage for England. The main weakness of this study is the lack of internal consistency in the use of Post-COVID syndrome codes, with a lack of correlation between the recording of diagnosis and referral codes. This lack of internal consistency in code usage means we cannot have confidence that any computed referral rate would be robust. Similarly, we cannot measure uptake of other recommendations in the guideline without an accurate denominator of applicable patients denoted by a diagnosis code.

The YCR website also offers post-discharge advice for those hospitalised with COVID-19 infection, so referrals to this source of self-help may include those who do not meet the criteria for Ongoing symptomatic COVID-19 or Post-COVID-19 syndrome. However, the levels of recording suggest that referrals to post covid clinics are often recorded without a formal Ongoing symptomatic COVID-19 or Post-COVID-19 diagnosis code. It is hard to evaluate to what extent low recording rates for the Your Covid Recovery website and programme reflect lack of signposting or lack of coding depth.

### Policy Implications and Interpretation

There are large numbers of referrals to Post-COVID-19 services without an associated diagnosis code being recorded. It is unclear why this is the case, but possible explanations include GPs not seeing a benefit to recording diagnosis and referral codes separately, perhaps using free text fields instead. The fact that referral codes are being used in greater numbers than diagnosis codes suggests that diagnosis codes are less likely to be recorded.

A failure to record a diagnosis code may have deleterious effects. Primary care diagnoses of Post-COVID syndrome need to be better recorded if we are to use EHR records to estimate Post-COVID syndrome prevalence or referral rates, or conduct future research using these sources. NICE’s “managing the long-term effects of COVID-19” guideline does not explicitly state diagnosis ought to be made in primary care. NHS England’s Enhanced service specification for Post-COVID syndrome required practices to “code consistently and accurately”, with codes specified in the Primary Care coding minimum dataset [12].

## Conclusion

This study demonstrated variation in diagnosis and referral codes for Post-COVID syndrome across different patient groups. The results, with limited crossover of referral and diagnostic codes, suggests the underuse of codes generally. This underuse limits the use of routine data for monitoring Post-COVID syndrome diagnosis and management, and compliance with best practice – but suggests several areas for improvement in coding and practice.

## Data Availability

All data in the manuscript has been extracted from the OpenSAFELY-EMIS-TPP platform. See https://www.opensafely.org/about/ for details.

## Appendix Long-term effects of COVID-19 SNOMED Diagnosis and Referral Codes

Appendix – Long-term effects of COVID-19 SNOMED Diagnosis and Referral Codes
| Code | Description |
| --- | --- |
| 1325181000000106 | Ongoing symptomatic disease caused by severe acute respiratory syndrome coronavirus 2 (disorder) |
| 1325161000000102 | Post-COVID-19 syndrome (disorder) |
| 1325031000000108 | Referral to post-COVID assessment clinic (procedure) |
| 1325041000000104 | Referral to Your COVID Recovery rehabilitation platform (procedure) |
| 1325021000000106 | Signposting to Your COVID Recovery (procedure) |

## Additional Information

### Information Governance and Ethical Approval

NHS England is the data controller for OpenSAFELY-EMIS and OpenSAFELY-TPP; EMIS and TPP are the data processors; all study authors using OpenSAFELY have the approval of NHS England. This implementation of OpenSAFELY is hosted within the EMIS and TPP environments which are accredited to the ISO 27001 information security standard and are NHS IG Toolkit compliant[13,14].

Patient data has been pseudonymised for analysis and linkage using industry standard cryptographic hashing techniques; all pseudonymised datasets transmitted for linkage onto OpenSAFELY are encrypted; access to the platform is via a virtual private network (VPN) connection, restricted to a small group of researchers; the researchers hold contracts with NHS England and only access the platform to initiate database queries and statistical models; all database activity is logged; only aggregate statistical outputs leave the platform environment following best practice for anonymisation of results such as statistical disclosure control for low cell counts[15].

The OpenSAFELY research platform adheres to the obligations of the UK General Data Protection Regulation (GDPR) and the Data Protection Act 2018. In March 2020, the Secretary of State for Health and Social Care used powers under the UK Health Service (Control of Patient Information) Regulations 2002 (COPI) to require organisations to process confidential patient information for the purposes of protecting public health, providing healthcare services to the public and monitoring and managing the COVID-19 outbreak and incidents of exposure; this sets aside the requirement for patient consent[16].

Taken together, these provide the legal bases to link patient datasets on the OpenSAFELY platform. GP practices, from which the primary care data are obtained, are required to share relevant health information to support the public health response to the pandemic, and have been informed of the OpenSAFELY analytics platform.

This study was supported by Cathy Hassell and Andrew Menzies-Gow as senior sponsors, and approved by the NICE Research Governance process.

## Acknowledgements

We are very grateful for all the support received from the EMIS and TPP Technical Operations teams throughout this work, and for generous assistance from the information governance and database teams at NHS England / NHSX.

This study was supported by the “Characterisation, determinants, mechanisms and consequences of the long-term effects of COVID-19: providing the evidence base for health care services (CONVALESCENCE)” programme, funded by NIHR (COV-LT-0009).

